# Use of nicotine replacement therapy to reduce children’s exposure to second-hand smoke in the home: Findings of a pilot randomised controlled trial conducted in Scotland

**DOI:** 10.64898/2026.08.13.26360348

**Authors:** Rachel O’Donnell, Karen Mather, Tracy Henderson, Laura Sinclair, Rebecca Howell, Nicola McMeekin, Sean Semple

**Affiliations:** Institute for Social Marketing and Health, Faculty of Health Sciences and Sport, University of Stirling, Pathfoot Building, Stirling, Scotland, FK9 4LA; NHS Lanarkshire Health Improvement Team, Beckford Street, Hamilton, Scotland, ML3 0TA; Health Economics and Health Technology Assessment, Clarice Pears Building, School of Health & Wellbeing, University of Glasgow, G12 8TB

**Keywords:** smoking in the home, second-hand smoke exposure, nicotine replacement therapy, pilot randomised controlled trial, inequalities

## Abstract

**Introduction:** Children’s exposure to second-hand tobacco smoke is a preventable global public health issue, yet there is no consensus on how best to support families to create a smoke-free home. This pilot randomised controlled trial tested the feasibility of use of free nicotine replacement therapy combined with telephone-delivered support to reduce children’s exposure to second-hand smoke in the home, and inform a future full-scale trial.

**Methods:** Parents/carers aged 18+ years, who smoke in the home and care for one or more children aged 0-16 years were recruited through existing initiatives and social media. Participants were randomised to either the intervention (Group A) or control (Group B) arm. Group A received free posted-to-home nicotine replacement therapy, alongside fortnightly telephone calls to support smoking abstinence in the home. Group B were signposted to the Scottish Government’s ‘Take it Right Outside’ website which provides interactive advice on creating a smoke-free home. To measure second-hand smoke levels, participants installed an air quality monitor in their living room for 7 days to measure fine particulate matter (PM_2.5_) at baseline and 12-week follow-up.

**Results:** Approximately one-quarter (n=27/100) of the intended sample size was recruited. Median PM_2.5_ concentrations reduced in both the intervention (-36μg/m^3^) and control (-16mg/m^3^) groups. Retention rates and adherence rates to nicotine replacement therapy were 70% and above, with no risks and/or safety concerns reported, suggesting this approach is feasible and acceptable to participants. The estimated cost of delivering this 12-week intervention was £244 per individual.

**Conclusions:** Although recruitment rates were insufficient to recommend progression to a larger trial to test effectiveness of this approach in Scotland, this study could inform trial development in other countries where smoking in the home is commonplace. Insights regarding the alignment of smoke-free home interventions with broader smoking cessation initiatives could inform future policy and public health approaches.

*What is already known on this topic:*

- Children’s exposure to second-hand tobacco smoke (SHS) is a preventable global public health issue, and whilst there is no consensus on how best to support families to create a smoke-free home, the use of nicotine replacement therapy (NRT) for temporary abstinence from smoking in the home has shown promise in UK settings.

*What this study adds:*

- Use of free posted-to-home NRT alongside behavioural support by telephone to reduce children’s exposure to second-hand smoke in the home is feasible in terms of retention rates, NRT adherence, andDthe practicalities of intervention delivery within an established National Health Service.
- Recruitment to the study was difficult and participation rates were insufficient to inform the development of a future trial to study intervention effectiveness, which likely reflects a Scotland-wide reduction in the proportion of individuals who smoke in the home in the presence of children, and perceived stigma in acknowledging this practice to healthcare staff.

*How this study might affect research, practice or policy:*

- The effectiveness of this approach should be evaluated in countries where smoking in the home is more prevalent, meaning that recruitment is potentially easier, and where NRT is licenced for use.
- Using NRT for temporary abstinence from smoking in the home creates conditions conducive to subsequent smoking cessation, highlighting the value of integrating smoke-free home interventions within wider tobacco control policies and public health strategies.

## Introduction

Exposure to second-hand tobacco smoke (SHS) during childhood is associated with a wide range of adverse health outcomes, including greater risk of developing respiratory illnesses such as asthma, bronchiolitis, croup and lower respiratory tract infections.^1^ Creating and maintaining a smoke-free home is therefore a key strategy for reducing children’s SHS exposure and may also encourage smoking cessation among parents and caregivers.^1,2^ In addition to these immediate health benefits, children raised in smoke-free homes are less likely to initiate smoking during adolescence.^3^ Because smoking prevalence remains substantially higher among socioeconomically disadvantaged populations, children living in these households continue to experience disproportionately high levels of SHS exposure.^4^

Although protecting children from SHS is a public health priority, Article 8 of the WHO Framework Convention on Tobacco Control (FCTC) does not provide specific recommendations for reducing SHS exposure within private homes. This contrasts with the comprehensive FCTC guidance on protecting people from SHS in public places, workplaces and public transport. The absence of guidance relating to the home is notable given that this is the principal environment in which children are exposed to SHS.^5^ Systematic review evidence has also highlighted uncertainty regarding the most effective approaches for supporting socioeconomically disadvantaged families to establish smoke-free homes.^6^ Qualitative syntheses suggest that parents and caregivers with sole responsibility for young children often face practical barriers to smoking outside rather than in the home, particularly where housing offers little or no private outdoor space.^4^ These challenges are reflected in Scotland, where marked socioeconomic inequalities in children’s SHS exposure persist. Recent Scottish Health Survey (SHeS) data show that 9% of children living in the most deprived areas are exposed to SHS in the home, compared with fewer than 1% of those living in the least deprived areas.^7^ Similar inequalities in children’s SHS exposure have been documented internationally, including in the USA,^8^ Australia,^9^ Germany,^10^ Spain,^11^ Denmark^12^ and Japan^13^. Although only 4% of children in Scotland were reported to be regularly exposed to SHS at home in 2023, this equates to more than 30,000 children experiencing avoidable exposure to a recognised carcinogen.^14^ Together, these findings reinforce the need for innovative, equitable interventions that better support disadvantaged families to establish and sustain smoke-free homes.

Research conducted in Scotland suggests that supporting people who smoke in disadvantaged communities to establish smoke-free homes is both feasible and acceptable. However, sustaining behavioural change remains difficult where opportunities to smoke outside are limited or where other household members and visitors continue to smoke indoors.^15,16^ One potential solution is the use of nicotine replacement therapy (NRT) to support temporary abstinence, enabling smokers to avoid smoking inside the home. Although free NRT is currently available in Scotland only for smoking cessation, National Institute for Health and Care Excellence (NICE) guidance also recommends its use to support temporary abstinence in indoor settings.^17^ Supporting this approach, a randomised controlled trial (RCT) in deprived communities in Nottingham, England evaluated a 12-week smoke-free homes intervention comprising behavioural support, NRT for temporary abstinence delivered during home visits, and personalised feedback from indoor air quality monitoring. Compared with usual care, the intervention significantly reduced both household SHS concentrations and the number of cigarettes smoked indoors,^18^ and was shown to be cost-effective.^19^ However, because the intervention incorporated multiple components, the independent contribution of NRT could not be determined.

To investigate the effects of NRT alone, two pilot studies were subsequently undertaken with parents and caregivers living in disadvantaged areas of Edinburgh, Scotland.^20–22^ In the first study^20,22^ qualitative interviews with parents (n=17) recruited through Early Years Centres (EYCs), Family Nurse Partnership (FNP) services and community pharmacies indicated strong support for using NRT to achieve temporary abstinence at home, with participants viewing it as preferable to indoor e-cigarette use. In the second phase, 32 parents received a home visit from a smoking adviser to discuss NRT options, after which 20 collected free NRT from community pharmacies for use over 12 weeks. Follow-up interviews described a range of positive self-reported outcomes, including the establishment of smoke-free homes, reductions of at least 50% in indoor smoking, and smoking cessation for a small number of participants. None of these outcomes were objectively assessed. Parents also self-reported financial benefits and spending more time with their children, often exceeding their own expectations of behaviour change. Some participants shared NRT with partners, supporting emerging recommendations for whole-household smoke-free home interventions.^23,24^ While pharmacy, EYC and FNP staff (n=13) were supportive of the intervention, they identified the multi-step process for obtaining NRT as a barrier to engagement.

The second pilot study,^21^ conducted during the COVID-19 pandemic, recruited 25 parents and caregivers, including grandparents. Participants obtained a 12-week supply of NRT directly from community pharmacies, removing the need for an initial home visit, while indoor air quality was measured at baseline and follow-up to objectively assess SHS exposure. Self-reported outcomes described during an optional qualitative end-of-study interview were mixed: three participants established smoke-free homes, one of whom subsequently quit smoking, six reduced indoor smoking by 50% or more, four reported no (sustained) reduction and one increased smoking in the home. Participants suggested that telephone-delivered behavioural support would improve NRT use and that posting NRT directly to participants’ homes could overcome barriers associated with mobility, lone parenting and the stigma of discussing smoking in person. These findings informed refinement of the intervention by incorporating ongoing behavioural support, simplifying NRT delivery and strengthening assessment of NRT adherence.

The present study reports a pilot RCT evaluating the feasibility of providing free NRT by post alongside telephone-delivered behavioural support to reduce children’s exposure to SHS in the home. The primary objective was to assess the feasibility of trial procedures, including recruitment, randomisation, retention, adherence, intervention delivery and data collection. A secondary objective was to evaluate methods for collecting objective measures of SHS exposure together with resource use and economic data to inform the design of a future definitive trial.

## Methods

### Study design

A full description of the methods is available in the study protocol paper.^25^ Supplementary file 1 summarises the design of this pilot RCT. Participants were randomised 1:1 to the intervention (Group A) or control (Group B) arm by a member of the University of Stirling research team. Where multiple family members participated, all were allocated to the same study arm.

Group A received 12 weeks of free NRT delivered by post, alongside fortnightly telephone support and resources to reduce children’s exposure to SHS. An NHS intervention delivery team member discussed NRT options by telephone using an information sheet developed from previous pilot work.^21^ Each call also addressed understanding of SHS harms, strategies to reduce smoking in the home (including engagement of other household members who smoke), and development of a personalised smoke-free home plan using theory-and evidence-based AFRESH materials developed through intervention mapping.^26^ Participants received their chosen NRT, personalised plan, AFRESH resources and NRT guidance by post. Product suitability was reviewed after one week, with alternative NRT provided where needed, followed by fortnightly supplies as needed and telephone reviews to support adherence and revised personalised plans if required.

Group B participants received current NHS Lanarkshire ‘standard care’ for those wishing to create a smoke-free home: a link to the Scottish Government’s *Take It Right Outside* website (https://www.nhsinform.scot/self-help-guides/second-hand-smoke-your-smoke-free-tips), providing interactive advice on creating a smoke-free home. At week 12, they were offered AFRESH resources, telephone advice on NRT options, and 12 weeks of free NRT posted-to-home fortnightly.

### Inclusion and exclusion criteria

Eligible participants were parents, carers, or relatives aged ≥18 years who smoked in the home and cared for at least one child aged 0–16 years for ≥1 day per week. Multiple family members could participate, and individuals were eligible if they lived with another adult who smoked and did not wish to participate. Although a completely smoke-free home may not have been achievable in these circumstances, reducing children’s exposure to SHS in the home was considered possible.

Individuals unable to understand or speak English as their primary language were excluded due to the absence of bilingual recruitment and intervention delivery resources. Exclusion criteria also included use of medications requiring medical monitoring alongside NRT use (see^25^ for full list), pregnancy or breastfeeding (as in this context available NRT products were licensed only for those making a quit attempt), and hypersensitivity to nicotine or NRT components.

### Setting

Lanarkshire has an estimated total population of 655,000 and an adult smoking prevalence rate of 14.7%, ranking 13th highest among Scotland’s 14 health board areas.^27^ NHS Lanarkshire has a specialist smoking cessation service (‘Quit your Way’) and over 100 community pharmacies providing behavioural support and cessation products. NHS Lanarkshire established a postal delivery service for NRT products through its Quit your Way Service, during the COVID-19 pandemic, based on practice in another Scottish health board area.

### Participants

Participants living in Lanarkshire were recruited by NHS health improvement team members between January 2024 and September 2025 through a wide range of strategies, including contact with third-sector organisations, voluntary groups and community settings. Recruitment also involved distribution of study flyers and information sheets at community venues and service centres, presentations to partner organisations, engagement with over 70 organisations by email, NHS digital promotion, targeted social media advertising, community events, employer engagement and promotion through all primary and secondary schools in Lanarkshire. Sample size was based on guidance suggesting a median of 36 participants per arm for pilot RCTs with similar primary outcomes.^28^ Allowing for planned qualitative interviews, sample characteristics and resources, the target recruitment was 100 participants, anticipating approximately 35 per group completing follow-up after up to 30% attrition.

### Measures

Following informed consent, a member of the NHS health improvement team contacted eligible individuals, who then completed a baseline telephone questionnaire assessing tobacco use, home smoking behaviours and heaviness of smoking using the Heaviness of Smoking Index. Deprivation postcode area was measured by the Scottish Index of Multiple Deprivation (SIMD).

Parents were offered the option of child saliva sampling and/or indoor air quality monitoring to assess children’s SHS exposure at baseline and 12-week follow up. Relatives providing childcare in their own homes were not asked to collect child saliva samples, as parental consent would be required, but they were eligible for air quality monitoring. Where parents cared for a single child aged under five years, air quality monitoring was conducted but saliva sampling was not offered, due to practical and safety considerations. Where a child aged ≥5 years was present, parents could choose saliva sampling and/or air quality monitoring. Saliva sampling procedures followed procedures used in the Health Survey for England (see^25^ for full details).

With permission, an NHS intervention delivery team member installed a PurpleAir Flex air quality monitor in the main living room of the home to measure fine particulate matter (PM_2.5)_ concentrations, a marker of SHS levels, in the home every two minutes for a period of one week. A follow-up telephone call was arranged during the visit to begin intervention delivery. At week 6, participants completed a process evaluation questionnaire assessing pilot-trial procedures, safety issues and intervention engagement. At week 12, follow-up data were collected on tobacco use, home smoking practices, heaviness of smoking, quit attempts, nicotine dependence, and safety concerns. Participants were invited to complete a qualitative interview at week 13 exploring intervention experiences, NRT use, adherence, changes in home smoking, consumption and smoking-related expenditure. Participants opting to quit smoking during the study were referred to NHS Lanarkshire’s Quit Your Way cessation service. Intervention participants who quit continued to receive fortnightly contact to support smoke-free home maintenance and NRT use where required.

The intervention delivery team recorded reflections on feasibility, delivery experiences and suggested process improvements. Resource data were collected to assess feasibility of a future cost-effectiveness analysis, following guidelines for pilot intervention studies.^29^ We conducted a literature review to inform a cost-consequences framework, and developed a resource use form to capture NRT type, dose, frequency and staff time involved in telephone support.

### Analysis

Recruitment, randomisation and retention outcomes were analysed descriptively to address feasibility objectives. Data included numbers screened, eligible and recruited; characteristics of non-consenting and ineligible participants; retention by trial arm; completion of 12-week follow ups; and reasons for withdrawal. Data were also collected on intervention engagement, including NRT receipt and adherence, use of personalised smoke-free home plans and participation in telephone support calls. Analysis of the objective measure of children’s exposure to SHS was also performed.

Progression criteria were developed with the Study Advisory Group, following current guidance,^29^ using a red/amber/green traffic light system^30^ to inform decisions regarding progression to a future definitive future trial (see Supplementary file 2).

### Ethics

Ethical approval was obtained from the West of Scotland Research Ethics Service 3 (15/12/2023, ref 23/WS/0153; amendment 13/12/2024, ref AM01) with Research and Development Management approval from NHS Lanarkshire (22/05/2024, L22083). The study protocol^25^ was registered with the ISRCTN registry (ISRCTN79307718). Participants received two £25 supermarket vouchers – one following completion of the week 6 process evaluation and one at study completion. Reporting followed the CONSORT extension for pilot trials guidelines (see Supplementary file 3).^31^

### Patient and Public Involvement

The study design was co-produced with individuals who smoke through two pilot studies conducted with mothers, fathers and caregivers living in low-income areas in Edinburgh, Scotland.^20–22^ Two public involvement groups also helped shape the development and focus of this study - the University of Nottingham Tobacco and Nicotine Group and an Edinburgh based Fathers group established by the lead author in 2018. Both groups felt fortnightly telephone calls in the intervention arm of the study would be beneficial and not too onerous, and they advised on reimbursement rates for study participants. Input from the Fathers group ensured our recruitment approaches/ materials were inclusive, which is important given smoke-free home interventions have tended to focus on the role of mothers and women in reducing children’s SHS exposure.^32^ We also discussed recruitment challenges with both public involvement groups, and their views shaped decision-making on increasing re-imbursement rates and reframing the wording of the recruitment flyer.

## Results

Recruitment was scheduled to take 18 months from September 2023 to March 2025. Due to a delay in set-up, recruitment began in June 2024, then proceeded to be consistently below the predicted target, and on this basis, the study team closed recruitment after 13 months, in September 2025. Supplementary file 4 shows the CONSORT flow diagram for the intervention (A) and control (B) groups.

Table 1 presents the sample characteristics for Group A and B populations. Four-hundred and sixty-two individuals expressed an initial interest in taking part, of which 47 were recruited to the study. Reasons for ineligibility included vaping rather than smoking in the home, and living with young adults aged 17 or older, rather than children aged 0-16 years. Eighty-three individuals expressed an interest in quitting smoking at first contact and were referred to the local Quit Your Way cessation service on that basis. Twenty-seven were successfully recruited to the study and randomised to either Group A (n=17) or Group B (n=10).

**Table 1:** Characteristics of study participants (n=27)

|  | Overall | Group A<br>(intervention) | Group B (standard<br>care) |
| --- | --- | --- | --- |
| Number of participants | 27 | 17 | 10 |
| SIMD<sup>#</sup>: decile mean (range) | 2.7<br>(1-6) | 2.6<br>(1-6) | 2.7<br>(1-5) |
| Gender (M/F) | 3/24 | 0/17 | 3/7 |
| Index child age mean (range) | 8.4<br>(0.5-16) | 8.3<br>(2-16) | 8.6<br>(0.5-15) |
| Heaviness of Smoking Index<br>score (category) | 3<br>(Medium) | 2<br>(Medium) | 3<br>(Medium) |
| Cigarettes smoked in the<br>home per day mean (range) | 11<br>(1-20) | 9<br>(1-20) | 13<br>(2-20) |
| Regular visitors smoke in the<br>home (yes/total) | 8/27 | 6/17 | 2/10 |
| Baseline PM<sub>2.5</sub> average: mean<br>(range) in µg/m<sup>3</sup> | 118<br>(3-426) | 143<br>(30-426) | 69.4<br>(3-149) |
| Baseline PM<sub>2.5</sub> peak<sup>^</sup>: mean<br>(range) in µg/m<sup>3</sup> | 1120<br>(94-2610) | 1090<br>(450-2610) | 1160<br>(94-2500) |
| Baseline PM<sub>2.5</sub> % time<br>>35 µg/m<sup>3</sup>: mean (range)* | 46.5<br>(0-97) | 52.1<br>(16-97) | 36.1<br>(0-76) |

<sup>#</sup> The Scottish Index for Multiple Deprivation decile (A score of 1 is the 10% most deprived; 10 is the 10% most affluent)
<sup>^</sup> The peak exposure refers to the highest 2-minute concentration recorded in the home.
\* The 35 µg/m<sup>3</sup> threshold is used as a marker of the proportion of time where the household PM<sub>2.5</sub> concentration exceeded the US EPA 'unhealthy for sensitive groups' guidance value for fine particulate pollution.

Baseline characteristics were broadly equivalent across Groups A and B. The mean age of the index child was eight years old in both groups, the mean deprivation decile was 2 in both groups, indicating that participants were generally from more deprived areas. The median number of cigarettes smoked per day was slightly higher in the control group (13) compared with the intervention group (9). Both groups consisted mainly of individuals with medium nicotine dependence.

### Feasibility of this approach

Approximately one-quarter (n=27/100) of the intended sample size was recruited. Over 70% (12/17) of those assigned to the intervention arm and 90% of those assigned to the control arm (9/10) completed the study process. All recruited and randomised study participants opted for household PM_2.5_ measurement, while only 15 agreed to provide a saliva sample from their child (data not analysed). Data on household PM_2.5_ concentrations at baseline was lost (device failure) for one of the participants in the control arm, and two of the participants in the control arm lived in the same home, meaning that baseline-follow-up paired data was available for a total of 20 participants from 19 homes (12/17 in the intervention arm and 9/10 in the control arm). In the intervention arm, over 70% (12/17) received more than one NRT delivery and engaged with at least one fortnightly telephone call. No risks and/or safety concerns associated with this approach were reported by participants. Pilot feasibility findings according to the pre-developed progression criteria are summarised in Table 2.

**Table 2:**
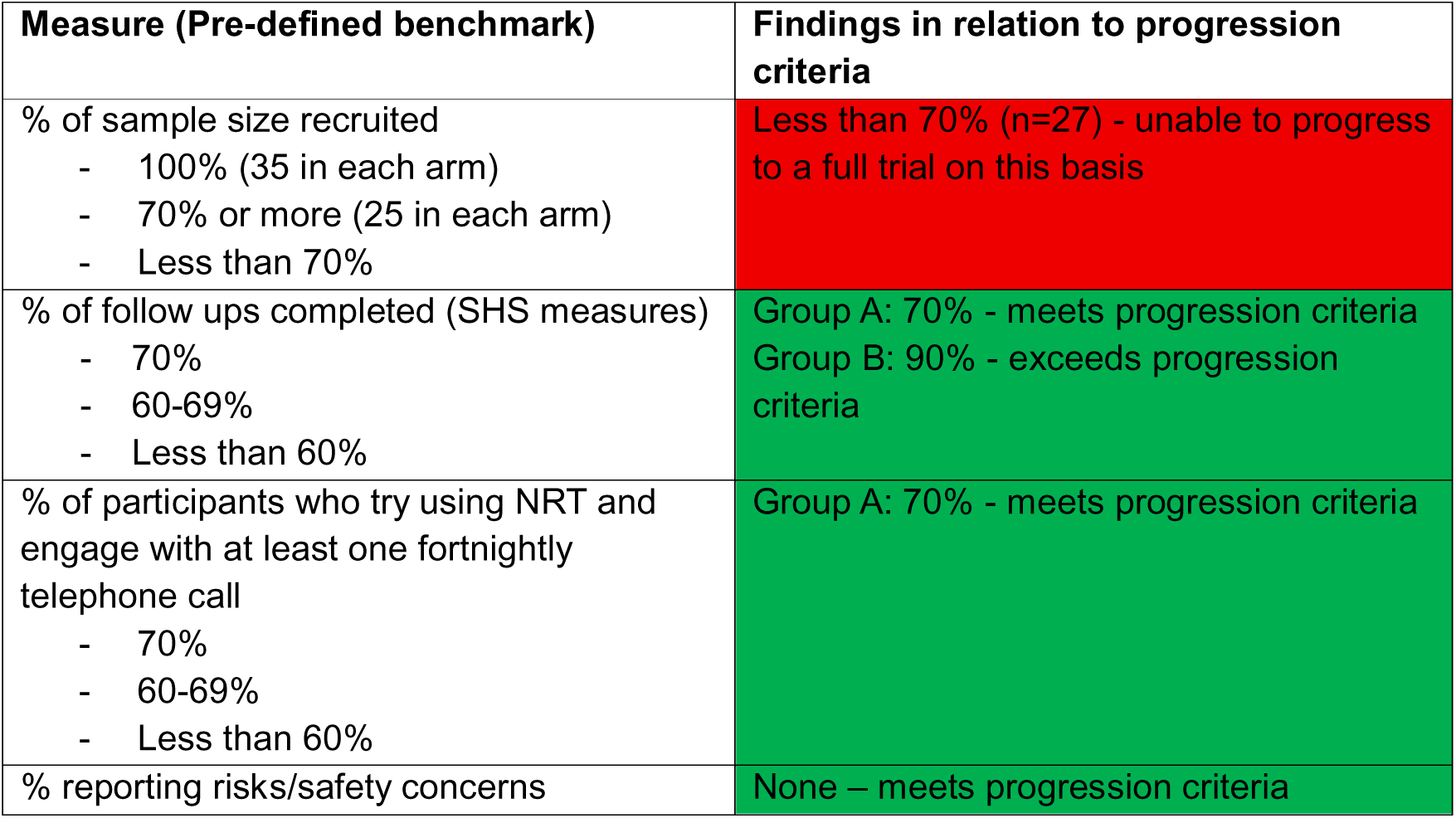
Pilot feasibility findings.

Detailed findings from the process evaluation and qualitative interviews conducted will be published separately. Supplementary file 5 provides a summary overview of NRT uptake and quantity posted across the 12-week intervention (as a proxy for engagement in at least 1 fortnightly call) – given these criteria were used to judge adherence to the intervention, alongside self-reported outcomes.

### Self-reported changes in household smoking

Follow up questionnaires were analysed for the 20 participants who had valid baseline and 12-week air quality monitoring. Not all participants provided a response to all questions. At 12-weeks follow-up ten out of twelve (83%) Group A homes reported becoming smoke-free (compared with two out of seven (29%) in Group B). Within the A group only two homes indicated cigarettes were smoked in the home at follow-up: one reducing from seven to three per day and the other increasing from seven to ten per day. Group A homes had a median (interquartile range) change in the number of cigarettes smoked in the home per day of-5 (-10 to-2.50) (Group B homes reported a similar change of-5 (-12 to 0)).

### Objective measurements of household air quality

The median (inter-quartile range) difference between baseline and follow-up PM_2.5_ measurements for Group A homes (n=12) was-36 (-70 to-25); Group B (n=7) was-16 (-47 to 162) μg/m^3^. A similar pattern was found when the change was expressed as a percentage change relative to the baseline measurement to account for the variation in measured concentrations at baseline. Figure 1 illustrates this change by paired measurements for each home with each data point providing the baseline and follow-up average PM_2.5_ concentrations measured. Given the small number of homes that completed air quality measurement we have not performed any statistical comparisons of the difference between the intervention and control group.

**Figure 1:**
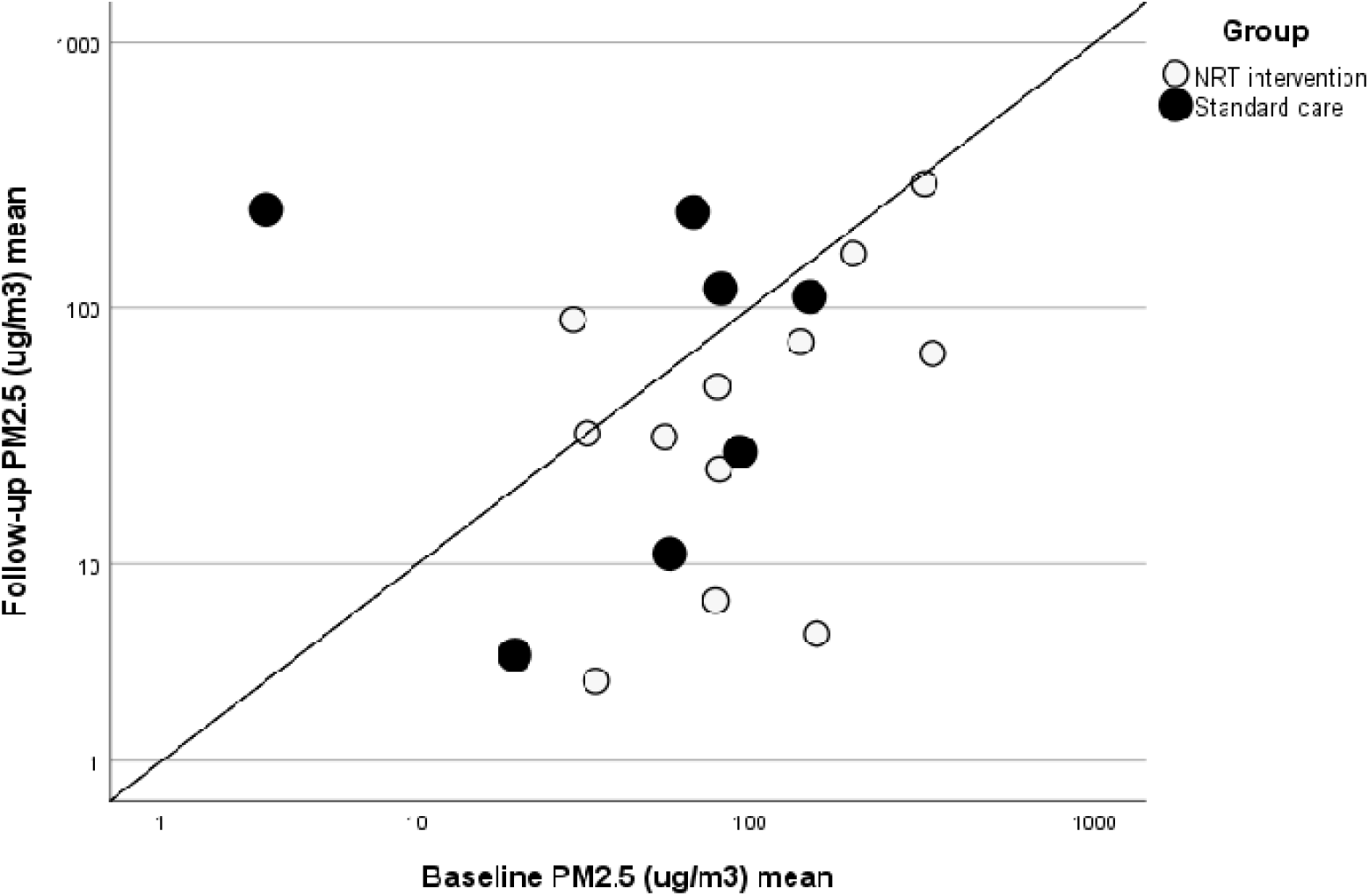
Scatterplot illustrating the paired PM_2.5_ average values from each home measured at baseline and then again at 12 week follow-up, divided by allocation group **Note:** A group = clear circles; B group = black circles. The black 1:1 line represents zero change; points to the left of the line indicate an increase in SHS levels after 12-weeks and points to the right of the line indicate homes that had reduced SHS levels after 12-weeks.

### Health economics data

Resource use data was available from all 17 participants randomised to Group A (the intervention arm), 12 of whom completed the intervention. There was no missing data. All Group A participants are included in this analysis (two participants withdrew at first support and did not receive any NRT products, however they received some support calls). The most popular NRT product requested was the inhalator (n=9), followed by mini lozenges (n=6), gum (n=3) and quickmist (n=2) (see Table 3).

**Table 3:** Health Economics Results.

| **Per Participant (n=17)** | **Mean** | **Minimum** | **Maximum** |
| --- | --- | --- | --- |
| **Nicotine Replacement Therapy** |  |  |  |
| Number of prescriptions | 3.5 | 0 | 7 |
| Number of products | 1.2 | 0 | 2 |
| Postage costs | £12 | £0 | £27 |
| NRT product costs | £155 | £0 | £499 |
| Advisor support |  |  |  |
| Cost of first contact | £36 | £2 | £119 |
| Number of supports received | 7.1 | 1 | 11 |
| Cost of support: |  |  |  |
| - Preparation | £13 | £0 | £38 |
| - Delivery | £27 | £3 | £67 |
| Total advisor support costs | £76 | £24 | £198 |
| Total intervention costs | £244 | £24 | £611 |

The mean number of NRT prescriptions and products used per Group A participant was 3.5 (minimum 0, maximum 7) and 1.2 (minimum 0, maximum 2). The mean postage costs for the NRT products and the mean NRT products costs themselves were £12 (minimum £0 where no NRT products were posted; maximum £27) and £155 (minimum £0 where no NRT products were posted, maximum £499) respectively.

The mean cost of the first contact with the advisor was £36 per participant (minimum £2, maximum £119). The mean number of supports received per participant was 7.1 (minimum 1, maximum 11). The cost of the support was £76 (minimum £24, maximum £198), broken down into preparation costing £13 (minimum £0, maximum £38) and delivery £27 (minimum £3, maximum £67) (Table 4). In total, the mean cost of the intervention per participant was £244 (minimum £24, maximum £611). The cost-consequence results (Table 5) illustrate the disaggregated cost data from Table 4 and relevant outcome data.

**Table 4:**
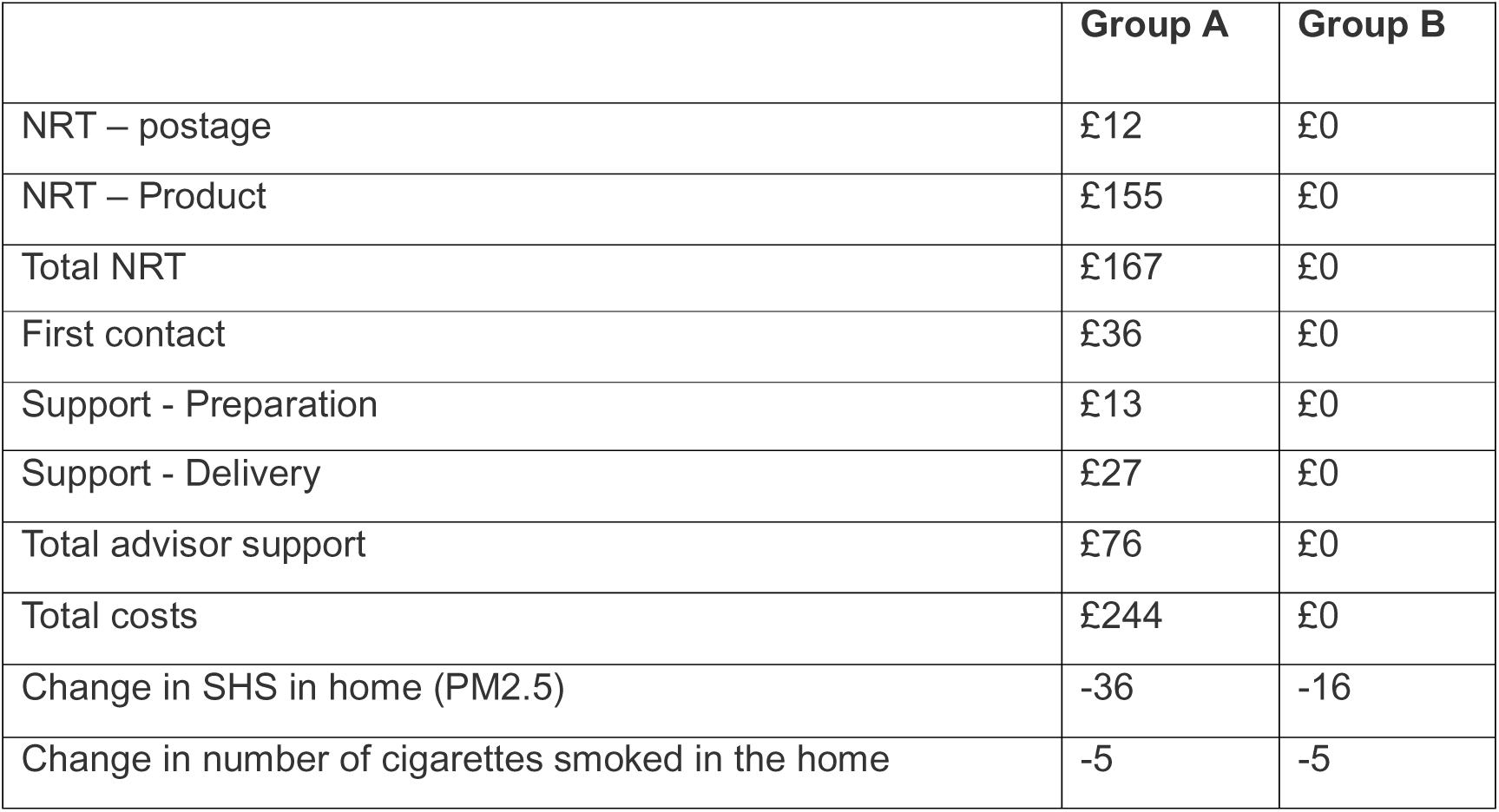
Cost-consequence analysis results.

|  | **Group A** | **Group B** |
| --- | --- | --- |
| NRT – postage | £12 | £0 |
| NRT – Product | £155 | £0 |
| Total NRT | £167 | £0 |
| First contact | £36 | £0 |
| Support - Preparation | £13 | £0 |
| Support - Delivery | £27 | £0 |
| Total advisor support | £76 | £0 |
| Total costs | £244 | £0 |
| Change in SHS in home (PM2.5) | -36 | -16 |
| Change in number of cigarettes smoked in the home | -5 | -5 |

The intervention resource data collected was suitable to estimate the intervention cost in a full trial and to inform a cost-consequence analysis. In a full trial we would also be interested in collecting healthcare (primary and secondary) resource use and potentially extrapolating the results to a lifetime horizon. Including this lifetime horizon would allow the long-term health harms/benefits of continued/reduced SHS exposure in the home to be captured, along with the increased likelihood of children who are exposed to SHS in the home becoming a smoker and the related subsequent smoking harms.

## Discussion

This study tested the feasibility of using free posted-to-home NRT alongside behavioural support by telephone to reduce children’s exposure to SHS in the home. Recruitment was challenging and insufficient participation rates suggest that a larger trial to study intervention effectiveness is impractical. The difficulties experienced in recruiting to this pilot likely reflect both a Scotland-wide reduction in the proportion of individuals who smoke in the home in the presence of children, and perceived stigma in acknowledging this practice to healthcare staff.^22^ Only one set of data were lost due to technical reasons. These findings build on previous pilot studies conducted in Scotland,^20–22^ and an RCT conducted in Nottingham, in England.^18,19^ Collectively, these studies show that this approach, which targets a socio-economically disadvantaged population who may find it particularly challenging to create a smoke-free home, is feasible, acceptable and cost-effective. This research also demonstrates several wider outcomes of this approach, including smoking reduction and associated financial benefits, and in some cases, smoking cessation.^18–21^

Associations between creating a smoke-free home and quit attempts, increased quit duration and reduced relapse have been emphasised in previous review findings.^33^ More recent prospective evidence similarly showed that adopting a smoke-free home increased quitting at both 3- and 6-month follow-up, and among those who continued smoking, was associated with fewer cigarettes smoked and more quit attempts.^2^ In our study, creating a smoke-free home appeared to’activate’ smoking cessation among individuals who initially had no intention to quit. Smoke-free home interventions may be viewed as less threatening than cessation-focused interventions,^2,34^ providing a more achievable initial goal that creates conditions conducive to subsequent smoking cessation.^2^

An unanticipated positive outcome of this study was that 83 individuals were referred to the local Quit Your Way cessation service after expressing motivation to quit during their first contact with the study team. This suggests that focusing on smoke-free homes, combined with broad recruitment, may engage individuals who are less likely to access conventional cessation services. Previous research indicates that disadvantaged groups are often underserved by cessation services because performance targets (e.g. four-week quit rates) do not align with the provision of the tailored, intensive support they may require.^35^ Our findings support suggestions that cessation service reach could be increased by starting to advise disadvantaged groups on smoke-free homes rather than directly with cessation.^35^ This bidirectional relationship highlights the value of integrating smoke-free home interventions within wider tobacco control strategies. For example, social housing providers are well placed to promote smoke-free homes and reduce smoking-related inequalities in underserved communities.^36^ Current NICE guidance recommending use of NRT for temporary abstinence provides a practical mechanism for integrating this approach into existing harm reduction and cessation services.^37^ This would broaden cessation service reach, and this approach could be well-utilised with individuals who engage with cessation services, struggle to quit successfully, but are still motivated to change their smoking behaviour. On this basis, and given collective study findings, we have developed a ‘How to Guide’ for NHS Health Boards in Scotland, should they wish to implement this approach at local level.^38^

Although recruitment rates were insufficient to recommend the development of a future, larger trial in Scotland to test effectiveness of this approach, our findings could be used as a basis for trial development in other countries where smoking in the home remains commonplace and NRT is licensed. This may be particularly relevant in South-East Asian countries, for example, where smoking prevalence and SHS exposure remain high, with the greatest burden falling on women and children.^39^ A high prevalence of multi-generational/multi-unit housing (MUH) in countries such as Malaysia and Singapore, and lower levels of awareness of the harmful effects of SHS, may mean that a randomised controlled trial is more viable within these countries. Residents of MUH are also more likely to include populations already at increased risk of chronic diseases and poor overall health, including those on low income, children, racial/ethnic minorities and older adults.^40^ While smoke-free housing policies and integrated cessation support have shown promise, particularly in the United States,^41^ non-smokers continue to be exposed to SHS because these approaches depend on effective cessation support, enforcement and voluntary compliance. These challenges highlight the need for interventions that support both smoking reduction and the creation of smoke-free homes, while carefully considering potential unintended consequences, including eviction risk, stigma and ethical concerns related to regulating smoking in private homes.^42^ Using NRT to support temporary abstinence in the home could address both objectives. Further research is therefore needed to determine whether the intervention tested in this pilot RCT could be adapted and implemented effectively in other countries.

In response to recruitment challenges, several protocol modifications were introduced. Participants were given the option of providing child saliva samples and/or home air quality measurements, as the requirement for saliva sampling had deterred some potential participants. Recruitment was expanded from the two most deprived SIMD quintiles to all quintiles to increase the eligible population. Following public involvement feedback, participant reimbursement was increased from a single £25 supermarket voucher at study completion to two £25 vouchers, one after the 6-week process evaluation and one at 12 weeks, consistent with a recent smoking cessation RCT.^43^ Recruitment materials were also revised to reduce potential stigma by reframing the invitation from “Do you smoke in the home?” to “Help us to help you create a smoke-free home.” Eligibility was broadened to include grandparents and other relatives providing regular childcare, and families with children under 5 years, although child saliva samples were not collected in this age group for safety reasons. These adaptations provide useful insights for the design of future smoke-free home trials. Future studies should also consider that some of the nicotine replacement therapy products used in this trial may not be routinely available in other settings.

A key strength of this study was its focus on intervention feasibility, as recruitment and retention challenges frequently undermine the success and policy impact of full-scale trials.^44^ Including parents, carers and relatives was another strength, reflecting the important caregiving role that extended family members often play in socioeconomically disadvantaged communities.^45^ However, the study also had limitations. Pregnant and breastfeeding women were excluded because the NRT products used are licensed only for those making a quit attempt. In addition, eligibility was restricted to individuals able to understand and speak English, limiting the generalisability of the findings and highlighting the need for culturally and linguistically adapted smoke-free home interventions for members of communities most affected by smoking, including migrant populations.^46^

## Conclusion

Providing disadvantaged households with free NRT for temporary abstinence alongside support for its effective use, may reduce children’s exposure to SHS in the home and offer an alternative pathway to smoking cessation. Although recruitment rates were insufficient to recommend the development of a future larger trial in Scotland, the study demonstrated acceptability of randomisation, feasibility of intervention delivery, high rates of retention and adherence to the intervention, and effective means of objective measurement of SHS exposure through PM_2.5_ measurement at baseline and follow-up. The effectiveness of this approach should be evaluated in countries where smoking in the home is more prevalent, meaning that recruitment is potentially easier, and where NRT is licenced for use. This study, and its protocol provide a foundation for future research in these international contexts.

## Data Availability

All data produced in the present study are available upon reasonable request to the authors

https://shine.ac.uk/nrt-for-sfhs-guide/

## Acknowledgements

We would like to thank both public involvement groups and participants from both previous pilot studies for helping to shape the research conducted, and our Study Advisory Group for their guidance and support. We would also like to thank Shirley Mawhinney for her valued support, guidance and input, and Dr Ruaraidh Dobson for early input into discussions regarding different ways to objectively measure children’s exposure to SHS in the home.

## Authors contributions

RO and SS conceived the idea for the study and RO is the guarantor. RO, SS, KM, TH, LS, RH and NM all contributed to the development of the pilot RCT. All authors contributed to protocol writing, and RO registered the protocol. RO led on writing this manuscript and all authors read and approved the final version.

