## Supplementary figures and images for "Use of nicotine replacement therapy to reduce children’s exposure to second-hand smoke in the home: Findings of a pilot randomised controlled trial conducted in Scotland"

### Supplementary file 1

**Supplementary file 1: Study design**

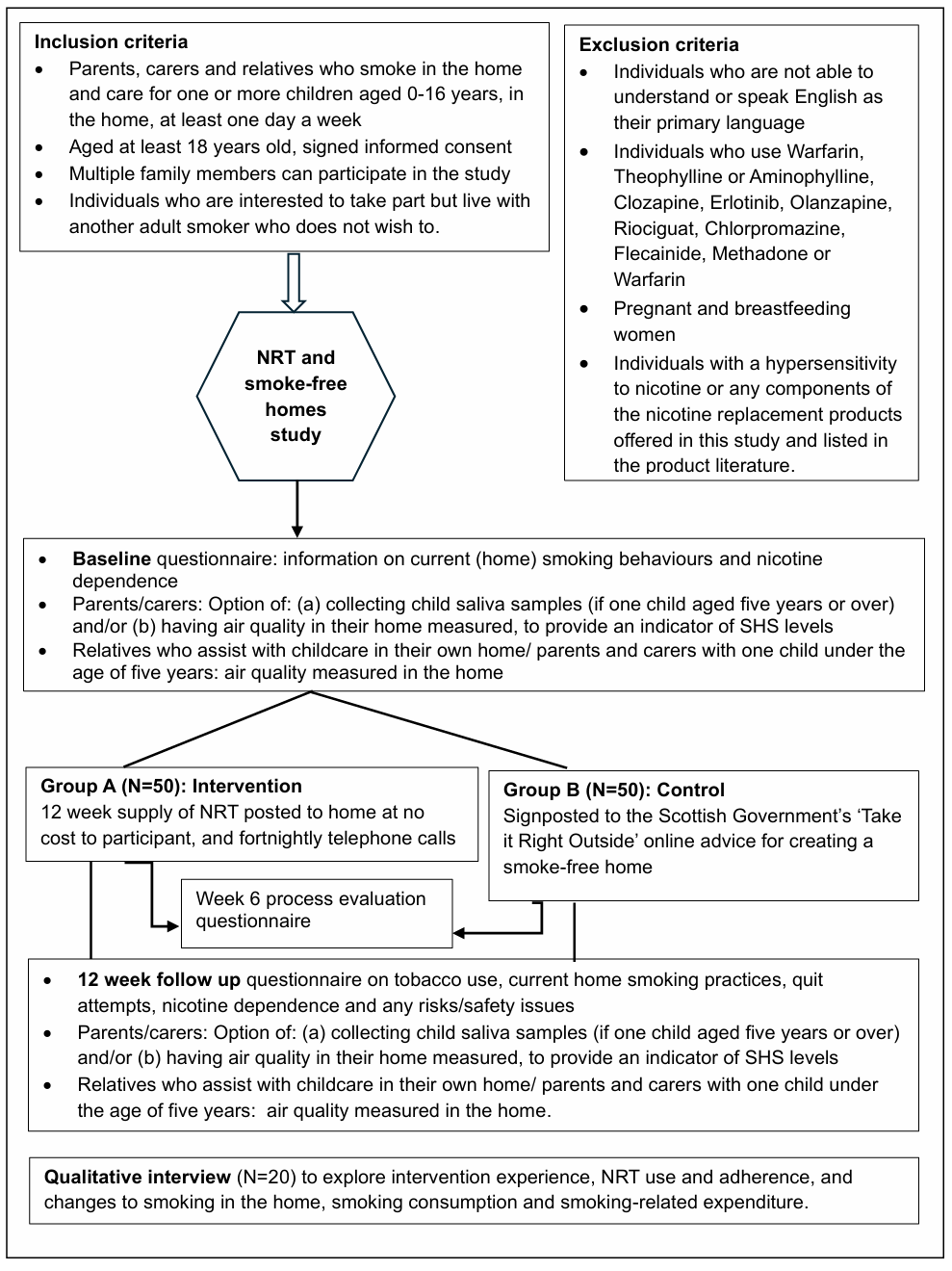
