## Supplementary file 2 for "Use of nicotine replacement therapy to reduce children’s exposure to second-hand smoke in the home: Findings of a pilot randomised controlled trial conducted in Scotland"

**Supplementary file 2: Developed progression criteria**

| **% of sample size recruited** | **100%**  **(35 in each arm)** | **70% or more**  **(25 in each arm)** | **Less than 70%** |
| --- | --- | --- | --- |
| **% of follow ups completed (SHS measures)** | **70%** | **60-69%** | **Less than 60%** |
| **% of participants who try using NRT and engage with at least one fortnightly telephone call** | **70%** | **60-69%** | **Less than 60%** |
| **% reporting risks/safety concerns** | **None** | **See note 1** | **See note 1** |

Note: The nature of reported risks and/or safety concerns could vary and include low, medium and high risks. On this basis, rather than specifying numbers for amber and red criteria, any reported risks/safety concerns will be discussed by the team and in conjunction with the Study Advisory Board to identify the extent to which they pose a concern in the context of decision making regarding a future definitive trial.
