## Supplementary file 3 for "Use of nicotine replacement therapy to reduce children’s exposure to second-hand smoke in the home: Findings of a pilot randomised controlled trial conducted in Scotland"

**Supplementary file 3: Completed CONSORT checklist**

| **Section/Topic** | **Item No** | **Checklist item** | **Reported on page No** |
| --- | --- | --- | --- |
| **Title and abstract** | | | |
|  | 1a | Identification as a pilot or feasibility randomised trial in the title | 1 |
|  | 1b | Structured summary of pilot trial design, methods, results, and conclusions (for specific guidance see CONSORT abstract extension for pilot trials) | 2 |
| **Introduction** | | | |
| Background and objectives | 2a | Scientific background and explanation of rationale for future definitive trial, and reasons for randomised pilot trial | 3-5 |
|  | 2b | Specific objectives or research questions for pilot trial | 5 |
| **Methods** | | | |
| Trial design | 3a | Description of pilot trial design (such as parallel, factorial) including allocation ratio | 5 |
|  | 3b | Important changes to methods after pilot trial commencement (such as eligibility criteria), with reasons | 12 |
| Participants | 4a | Eligibility criteria for participants | 6 |
|  | 4b | Settings and locations where the data were collected | 6 |
|  | 4c | How participants were identified and consented | 6 |
| Interventions | 5 | The interventions for each group with sufficient details to allow replication, including how and when they were actually administered | 5, 7 |
| Outcomes | 6a | Completely defined prespecified assessments or measurements to address each pilot trial objective specified in 2b, including how and when they were assessed | 7 |
|  | 6b | Any changes to pilot trial assessments or measurements after the pilot trial commenced, with reasons | 12 |
|  | 6c | If applicable, prespecified criteria used to judge whether, or how, to proceed with future definitive trial | 8 |
| Sample size | 7a | Rationale for numbers in the pilot trial | 6 |
|  | 7b | When applicable, explanation of any interim analyses and stopping guidelines | N/A |
| Randomisation: |  |  |  |
| Sequence  generation | 8a | Method used to generate the random allocation sequence | 5 |
|  | 8b | Type of randomisation(s); details of any restriction (such as blocking and block size) | 5 |
| Allocation  concealment  mechanism | 9 | Mechanism used to implement the random allocation sequence (such as sequentially numbered containers), describing any steps taken to conceal the sequence until interventions were assigned | 5 |
| Implementation | 10 | Who generated the random allocation sequence, who enrolled participants, and who assigned participants to interventions | 5 |
| Blinding | 11a | If done, who was blinded after assignment to interventions (for example, participants, care providers, those assessing outcomes) and how | N/A |
|  | 11b | If relevant, description of the similarity of interventions | N/A |
| Statistical methods | 12 | Methods used to address each pilot trial objective whether qualitative or quantitative | 8 |
| **Results** | | | |
| Participant flow (a diagram is strongly recommended) | 13a | For each group, the numbers of participants who were approached and/or assessed for eligibility, randomly assigned, received intended treatment, and were assessed for each objective | 8 and Suppl. file 4 |
|  | 13b | For each group, losses and exclusions after randomisation, together with reasons | 9 |
| Recruitment | 14a | Dates defining the periods of recruitment and follow-up | 8 |
|  | 14b | Why the pilot trial ended or was stopped | 8 |
| Baseline data | 15 | A table showing baseline demographic and clinical characteristics for each group | 8 and Table 1 |
| Numbers analysed | 16 | For each objective, number of participants (denominator) included in each analysis. If relevant, these numbers  should be by randomised group | 9 |
| Outcomes and estimation | 17 | For each objective, results including expressions of uncertainty (such as 95% confidence interval) for any estimates. If relevant, these results should be by randomised group | 9 and Table 2 |
| Ancillary analyses | 18 | Results of any other analyses performed that could be used to inform the future definitive trial | 9-10, Tables 3-4, Figure 1 |
| Harms | 19 | All important harms or unintended effects in each group (for specific guidance see CONSORT for harms) | 9 and Suppl. file 5 |
|  | 19a | If relevant, other important unintended consequences | 8 |
| **Discussion** | | | |
| Limitations | 20 | Pilot trial limitations, addressing sources of potential bias and remaining uncertainty about feasibility | 11-12 |
| Generalisability | 21 | Generalisability (applicability) of pilot trial methods and findings to future definitive trial and other studies | 10-12 |
| Interpretation | 22 | Interpretation consistent with pilot trial objectives and findings, balancing potential benefits and harms, and  considering other relevant evidence | 10-12 |
|  | 22a | Implications for progression from pilot to future definitive trial, including any proposed amendments | 10-12 |
| **Other information** | | |  |
| Registration | 23 | Registration number for pilot trial and name of trial registry | 8 |
| Protocol | 24 | Where the pilot trial protocol can be accessed, if available | 8 |
| Funding | 25 | Sources of funding and other support (such as supply of drugs), role of funders | 13 |
|  | 26 | Ethical approval or approval by research review committee, confirmed with reference number | 8 |
