## Supplementary file 4 for "Use of nicotine replacement therapy to reduce children’s exposure to second-hand smoke in the home: Findings of a pilot randomised controlled trial conducted in Scotland"

**Supplementary file 4: CONSORT flow diagram for the intervention (A) and control (B) groups**

Excluded (n=415)

¨  Not meeting inclusion criteria (n=29)

¨  Declined to participate (n=1)

¨  Referred to cessation service (n=83)

Other reasons (n=302)

Allocated to Group B (control) (n=10)

¨ Received allocated intervention (n=9)

¨ Did not receive allocated intervention (withdrew prior to baseline visit) (n=1 )

Lost to follow-up (withdrew after baseline visit) (n=3)

Analysed (n= 12) ¨ Excluded from analysis (n=0)

Analysed (n=7)
¨ Excluded from analysis (n=2): (baseline PM2.5 measurement failed) (n= 1); (two participants lived in same home) (n=1)

Lost to follow-up (give reasons) (n=0)

Discontinued intervention (give reasons) (n=0)

Assessed for eligibility (n=464)

### Follow-Up

### Analysis

Allocated to Group A (intervention) (n=17)

¨ Received allocated intervention (n=15)

¨ Did not receive allocated intervention (withdrew prior to visit) (n=2)

### Allocation

Randomized (n= 27)

### Enrolment
