## Supplementary file 5 for "Use of nicotine replacement therapy to reduce children’s exposure to second-hand smoke in the home: Findings of a pilot randomised controlled trial conducted in Scotland"

**Supplementary file 5: Intervention arm: NRT uptake, quantity posted and self-reported outcomes**

| **Participant ID** | **House type** | **NRT product choice/quantity posted across the 12 weeks** | **Self-reported baseline smoking behaviours** | **Self-reported outcomes** |
| --- | --- | --- | --- | --- |
| 02NRTSFH | Terraced House | Gum 2mg x 9 packs | Smokes in the kitchen  Smokes 11-20 per day (10 in the home) | **Smoke-free home**  Successful quit attempt |
| 03NRTSFH | Flat (own entrance) | Inhalator 36 cartridge x 3  Quick Mist Single 1mg x 1  Quick Mist Duo x 3 | Smokes in downstairs rooms  Smokes 11-20 per day (10 in the home) | **Smoke-free home**  Smoking 10 or less cigarettes per day outside |
| 04NRTSFH | Terraced House | Mini lozenges 2mg x 3 (60’s)  Mini lozenges 2mg x 3 (100’s) | Smokes in the kitchen  Smokes 11-20 per day (5 in the home) | **Smoke-free home**  Smoking 10 or less cigarettes per day outside |
| 06NRTSFH |  | Withdrew after randomisation | | |
| 09NRTSFH |  | Withdrew after randomisation | | |
| 16NRTSFH | Terraced House | Inhalator 36 cartridge x 7 | Smokes in all downstairs rooms  Smokes 11-20 per day (7 in the home) | Smoking allowed in kitchen/utility room  Smoking 10 or less per day (<5 in the home) |
| 21NRTSFH | Withdrew after baseline | | | |
| 22NRTSFH | Withdrew after baseline | | | |
| 24NRTSFH | Ground floor Flat | Mini lozenges 2mg x 6 (100’s) | Smokes in the kitchen and living room  Smokes 11-20 per day (15 in the home) | **Smoke-free home**  Smoking 10 or less cigarettes per day outside |
| 27NRTSFH | Terraced House | Inhalator 36 cartridge x 13 | Smokes in the kitchen  Smokes 11-20 per day (10 in the home) | **Smoke-free home**  Smoking 10 or less cigarettes per day outside |
| 29NRTSFH | Withdrew after baseline | | | |
| 32NRTSFH | Terraced house | Gum 2mg x 3 packs  Inhalator 36 cartridge x 2 | Smokes in the living room  Smokes 10 or less in per day (5 in the home) | **Smoke-free home**  Smoking 10 or less cigarettes per day |
| 33NRTSFH | Flat – above ground level | Mini lozenges 2mg x 1 (60’s)  Mini lozenges 2mg x 2 (100’s) | Smokes in the kitchen  Smokes 10 or less cigarettes per day (2 in the home) | **Smoke-free home**  Successful quit attempt |
| 34NRTSFH | Terraced house | Inhalator 36 cartridge x 11 | Smokes in the living room and bedroom  Smokes 11-20 cigarettes per day (18 in the home) | **Smoke-free home**  Unsuccessful quit attempt, smokes 11-20 cigarettes per day outside |
| 45NRTSFH | Flat – above ground level | Mini lozenges 2mg x 5 (60’s) | Smokes in the kitchen and bathroom  Smokes 10 or less cigarettes per day (5 in the home) | **Smoke-free home**  Successful quit attempt |
| 46NRTSFH | Terraced house | Inhalator 36 cartridge x 2  Mini lozenges 2mg x 1 (60’s) | Smokes in the kitchen/at the back door  Smokes 11-20 cigarettes per day (7 in the home) | Smoking at the back door  Smoking 10 or less cigarettes per day (5 at the back door) |
| 47NRTSFH | Flat – above ground level | Inhalator 36 cartridge x 14 | Smokes in the kitchen/living room and dining room  Smokes 10 or less cigarettes per day (1 in the home) | **Smoke-free home**  Successful quit attempt |
